# Graph-based Fusion Modeling and Explanation for Disease Trajectory Prediction

**DOI:** 10.1101/2022.10.25.22281469

**Authors:** Amara Tariq, Siyi Tang, Hifza Sakhi, Leo Anthony Celi, Janice M. Newsome, Daniel L. Rubin, Hari Trivedi, Judy Wawira Gichoy, Bhavik Patel, Imon Banerjee

## Abstract

We propose a relational graph to incorporate clinical similarity between patients while building personalized clinical event predictors with a focus on hospitalized COVID-19 patients. Our graph formation process fuses heterogeneous data, i.e., chest X-rays as node features and non-imaging EHR for edge formation. While node represents a snap-shot in time for a single patient, weighted edge structure encodes complex clinical patterns among patients. While age and gender have been used in the past for patient graph formation, our method incorporates complex clinical history while avoiding manual feature selection. The model learns from the patient’s own data as well as patterns among clinically-similar patients. Our visualization study investigates the effects of ‘neighborhood’ of a node on its predictiveness and showcases the model’s tendency to focus on edge-connected patients with highly suggestive clinical features common with the node. The proposed model generalizes well by allowing edge formation process to adapt to an external cohort.

## Introduction

Ability to automatically leverage patient similarity via a network architecture can improve efficiency in the healthcare system and contribute to the broader goal of precision medicine. Clinical event prediction is one of the key applications of big data and artificial intelligence, and instead of one-size-fits-all approaches, outcome prediction and decision making based on personalized models could be more accurate when patient similarity analysis has been taken into account. Existing literature depends on static similarity formulation based on pre-defined knowledge for predictive analysis without considering learned representation^1^.

Recent developments in graph convolutional neural networks (GCNN) have opened up the opportunity to learn effective representations for similarities between cases in addition to multi-modal data fusion by representing various modalities as edges and nodes of graphs. Availability of comprehensive public databases such as TADPOLE^2^, Alzheimer’s Disease Neuroimaging Initiative (ADNI)^1^ and ABIDE^2^ containing imaging and non-imaging information for patients with Alzheimer and autism spectrum disorder (ASD), respectively, has facilitated the use of GCN for disease prediction^3–7^ with innovations in GCN architecture involving kernel size selection and use of recurrence. We build on this research trend and build graphs with imaging data, i.e., X-rays as nodes, and weighted edges based on similarity in patients’ medical history recorded in EMR. We propose a novel framework for fine-grained clinical event prediction for COVID-19 patients by processing our patient-similarity graph through a graph convolutional neural network (GCN).

Much of the literature regarding predictive modeling for COVID-19 patients has been largely focused on either diagnosis by processing imaging data (chest X-rays) through deep convolutional neural networks (CNN)^8–13^, or mortality prediction using clinical risk factors^14–18^. In contrast, we propose a comprehensive relational graph approach to fuse information from imaging and non-imaging modalities. Furthermore, we propose a framework for multi-stage predictive modeling of disease trajectory of patients hospitalized with positive diagnosis of COVID-19, in terms of clinical events of discharge from hospital and mortality (Figure 01). While a pre-trained model was used for chest X-ray featurization, we developed innovative feature engineering schemes to model sparse information regarding past medical procedures and diagnosed illnesses of patients. Our relational graph-based modeling allows prediction processes to gather cues from similar cases, i.e., patients with similar demographic features and medical history. Our experiments indicate that our model that fuses imaging and non-imaging data through graph based modeling outperforms models using only imaging or only non-imaging EMR data. Graph based models do not require consistency in the graph formation process to ensure compatibility of the new graph with the trained model. This allowed us to update the clinical characteristics of patients used for formation of edges while validating our model on external data. Models trained on internal data were still compatible and performed reasonably well.

As deep learning models are generally opaque and it is a challenge to explain their decisions, we particularly focused on visualization of our models’ results. We designed a dashboard interface for exploration of model decision in clinical setting by drawing panels showing node features of the targeted case (chest X-ray image and relevant demographic features) as well as highlight the characteristics of ‘learned’ similar cases which is the ‘neighborhood’ of the target node, i.e., edge-connected nodes. We performed these visualization experiments for both internal and external sets. Varying neighborhood characteristics for the two sets corresponding to prediction of the same labels also highlight the unique adaptability of the graph-based modeling.

## Materials and Methods

We developed a graph-based fusion model to predict COVID-19 disease trajectory by using multi-modal patient data including chest X-ray imaging data, demographic information, and patient’s clinical history.

### Cohort description

Following approval of the Emory Institutional Review Board (IRB), we collected all chest X-rays of patients with at least one positive RT-PCR test for COVID-19, performed in 12 centers of the Emory Healthcare network from January 2020 to December 2020. As shown in Figure 1, the data consisted of 47,555 chest X-rays belonging to 23,831 unique patients. We only considered posteroanterior (PA) and anteroposterior (A) view of chest radiographs during the period of hospitalization. We also collected hospital admission, mortality and discharge data from the hospital billing system. Patients who were never admitted, or those who were admitted but did not receive rays at regular intervals (at maximum every three days) were discarded. This left 2,201 unique patients corresponding to 2,983 hospitalizations for which chest-x rays were obtained at least every 3 days (7,813 total chest-x rays). In order to avoid data leakage, we applied a patient-wise split of the 80% train and 20% test sets. Some patients were hospitalized multiple times during the year 2020. We considered the chest X-ray study date as a time point for predicting future clinical events and stratified these events based on severity – discharged from hospital within 3 days, or death. There were 929 patients and 4,850 chest X-rays after which the patient remained in the hospital for more than three days. There were 1,754 patients corresponding to 2,963 chest X-rays who were discharged from hospital within 3 days. 220 patients corresponding to 502 chest X-rays died within 3 days. Outcome data for this cohort was collected from warehouse EHR data sources. Our goal was to predict adverse events as early as possible to provide enough decision making time for clinicians. In our preliminary experiments, we evaluated 3 days up to 7 days, however given the unbalanced dataset, we found 3 days to be the most prevalent in terms of different prediction labels. This cut-off provided a rather balanced distribution of discharged vs. not discharged labels.

**Figure 1.**
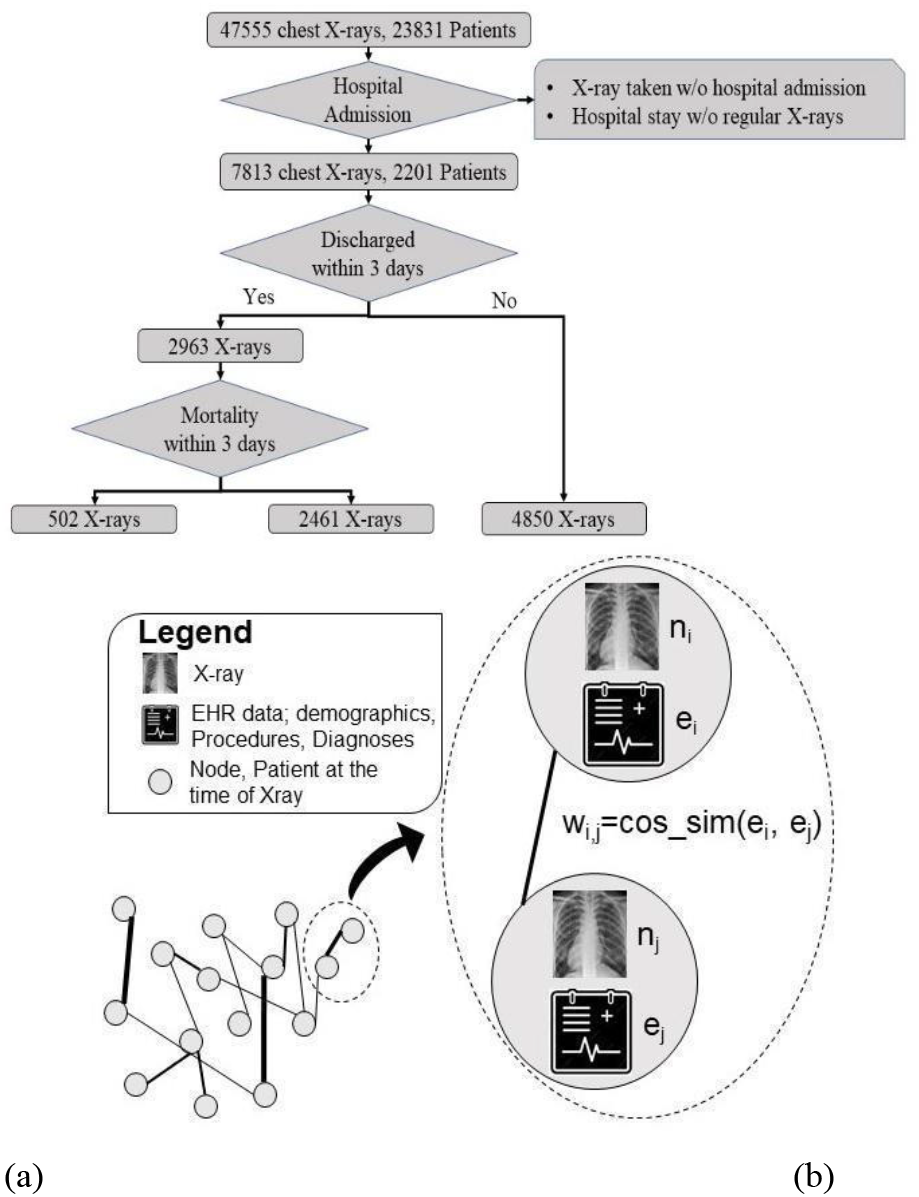
**(a)** CONSORT style diagram for internal cohort selection for training; **(b)** Proposed formation of graph with weighted edges for disease trajectory prediction

#### External test

With the approval of Mayo Institutional Review Board, we shipped our Emory trained model to Mayo and evaluated externally on a small set of 50 unique patients admitted to the Mayo Clinic hospital between Jan 2020 – Dec, 2020 with a positive RT-PCR test. The patients have 293 chest X-rays during the period of hospitalization. Demographic and comorbidities statistics of both cohorts are provided in Table 1.

**Table 1.** Study cohort characteristics - demographics, clinical history and insurance status. The numbers have been represented in terms of complete dataset (total cohort) as well as train and test split

| Characterization |  | Total cohort<br>(2201) | Train (1762) | Test<br>(439) | External<br>test (50) |
| --- | --- | --- | --- | --- | --- |
| Gender | Male | 1111 | 887 | 224 | 34 |
|  | Mean Age (std. deviation) | 59.7(16.2) | 59.2 (16.1) | 61.5(16.3) | 64.5 (16.3) |
|  | Female | 1090 | 875 | 215 | 16 |
|  | Mean Age (std. deviation) | 59.0 (18.8) | 58.7(18.9) | 60.2(18.4) | 59.8 (15.9) |
| Race | African American | 1405(63.8%) | 1135(64.4%) | 270(61.5%) | 6 (12%) |
|  | Caucasian | 554(25.2%) | 427(24.2%) | 127(28.9%) | 43 (86%) |
|  | Asian | 51(2.3%) | 42(2.4%) | 9(2.1%) | 1(2%) |
|  | American Indian or Alaskan | 11(0.5%) | 7(0.4%) | 4(0.9%) | -- |
|  | Multiple | 6(0.3%) | 6(0.3%) | 0(0%) | -- |
|  | Native Hawaiian or Other | 6 (0.3%) | 4(0.2%) | 2(0.4%) | -- |
|  | Unknown | 168(7.6%) | 141(8.0%) | 27(6.2%) | -- |
| Ethnicity | Hispanic | 163(7.4%) | 133(7.5%) | 30 (6.8%) | -- |
|  | Non-Hispanic | 1894(86.1%) | 1517(86.1%) | 377(85.9%) | 50 (100%) |
|  | Unknown | 144(6.5%) | 112(6.4%) | 32(7.3%) | -- |
| Comorbidities | Diabetes | 1187(53.9%) | 937 (53.2%) | 250(56.9%) | 7(14%) |
|  | Renal Disease | 1330(60.4%) | 1050(59.6%) | 280(63.8%) | 10(20%) |
|  | Hypertension | 1711(77.7%) | 1356(77.0%) | 355(80.9%) | 19 (38%) |
|  | Respiratory Disease | 1948(88.5%) | 1556(88.3%) | 392(89.3%) | 27 (54%) |
| Insurance Status | Medicare | 1034(47.0%) | 806(45.7%) | 228(51.9%) | Not available |
|  | Commercial | 784(35.6%) | 651 (36.9%) | 133(30.1%) |  |
|  | Medicaid | 125(5.7%) | 100(5.7%) | 25(5.7%) |  |
|  | Others | 258 (11.7%) | 205(11.6%) | 53(12.1%) |  |
| Alcohol Use | No | 1464(66.5%) | 1178(66.9%) | 286(65.2%) |  |
|  | Yes | 245(11.1%) | 213(12.1%) | 32(7.3%) |  |
|  | Not Reported | 492(22.4%) | 371 (21.1%) | 121(27.6%) |  |

### Graph-based Fusion model - Image and EHR

GCN model learns vector embeddings of nodes optimized for a down-stream prediction task. Difference between convolutional neural networks and GCN comes from the definition of neighborhood. CNN allows only spatial proximity to define the neighborhood that may have influence over the representation learning for an instance. GCN leaves the definition of neighborhood upto the designer and receives this information in the form of edges of a graph. Edge-connected nodes may influence the representation learning for a node. Such flexibility is particularly suited to model complex clinical scenarios. In our graph design, we used the chest X-ray embeddings as nodes and edges are decided by similarity in patients’ demographic information or medical history (see Figure 2b). Therefore, our GCN supports the notion of ‘neighborhoods’ based on clinical and/or demographic similarity between patients and makes predictions for future clinical events by considering trends among similar patients.

#### Graph node formation

We used a DenseNet-121^19^ pre-trained on the open-source CheXpert chest X-ray dataset^20^, and fine-tuned on 199,029 non-COVID chest X-rays from EUH acquired in 2019, for processing normalized images. We dropped the final softmax classification layer of the DenseNet-121 model and extracted 1024-dimensional feature vectors from each image to construct a dense representation of the images for the graph node formulation.

#### Similarity based edge formation

We encoded EMR features of the patient corresponding to a node in a binary vector using only clinical data collected before the study datetime of the chest X-ray to avoid any data leakage. We applied three edge design styles based on different clinical factors - demographic, procedure and comorbidities. Cosine similarity between EMR feature vectors corresponding to two nodes is used to decide the edge between two nodes, while edge weight encodes the strength of the similarity between those nodes.

- *Demographic information:* We used one-hot encoding for the following demographic features; gender (male/female), self-reported race (African American, Caucasian, Native Hawaiian or Other Pacific Islander, Asian, American Indian or Alaska Native, Multiple, Unknown), ethnic group (Hispanic or Latino, Non-Hispanic or Latino, Unknown), age at the time of admission (binned in 10-year intervals).
- *Current Procedural Terminology codes (CPT):* CPT is a five-digit procedure code that reports medical, surgical, and diagnostic procedures and services to entities such as physicians, health insurance companies and accreditation organizations. CPT codes are maintained and grouped in a hierarchical structure by the American Medical Association (AMA). Each CPT code was reduced into a higher-order parent category based on the defined hierarchy ^3^. We selected 21 groups with more than 1000 occurrences.
- *Comorbiditie*s: Past and current diagnoses of patients are structured as International Classification of Disease, 9th edition (ICD-9) codes which are grouped based on hierarchical structure^21^. We selected 29 groups occurring more than 5000 times in the data.

Many GCN variations are designed for transductive learning such that they can only process graph structures used for training with limited ability to generalize to unseen nodes or new graph structures. To avoid this limitation, we used the SAGE (SAmple and aggreGatE) graph convolution network (GraphSAGE)^22^ that optimizes a sample aggregate function to collect ‘messages’ from neighboring nodes while generating vector embedding of a node. For inference, GraphSAGE employs an optimized aggregate function to generate embedding for unseen nodes in unseen graph structures. GraphSAGE based prediction models can inductively reason to assign predictive labels to unlabeled nodes by learning from labeled nodes in the graph.

### Branched Framework of Prediction

Our work is focused on forecasting the trajectory of disease, in terms of two clinical events, once the patient has been hospitalized. Given the challenge of collecting a balanced dataset for multi-class classification, we modeled 2 sequential decision points and developed a pipeline for comprehensive prediction of possible clinical events. (Model1) - Prediction of discharge from the hospital: In the first decision point, Model-1 predicts whether the patient will stay in the hospital for more than 3 days (positive label) or not. (Model-2) - Mortality prediction: For negatively labeled instances by Model-1, in the second decision point, Model-2 predicts whether the patient will expire within 3 days (positive label) or not. Distribution of positive and negative class labels is highly imbalanced, especially for Model-2 (Figure 2). We employed undersampling of majority labels and weighted loss to tackle this challenge. A patient is evaluated every time a chest radiograph is taken, while staying in the hospital. Disease trajectory can be indicated as improving (discharge in less than 3 days) or worsening (mortality in 3 days).

### Model comparison and statistical evaluation

Evaluation of the proposed modeling framework is focused on two aspects; 1) performance of the fusion graph-based model in comparison to traditional modeling using single modality (either imaging data or non-imaging data), 2) comparative effectiveness of different EMR data sources in terms of graph structure definition to identify optimal similarity metrics for the targeted prediction task. We designed a Random Forest classifier and used the Pulmonary X-ray severity (PXS) scores computed by a CNN^23^ to predict clinical events as well. As PXS scores are based on deep learning based processing of imaging data, we wanted to establish the benefits of our selection of imaging features in our relational graph by comparison. We report the model performance in terms of Area Under the Receiver Operating Characteristics curve (AUCROC), precision, recall and f1-score on held-out test-set from Emory University cohort, as well as on an external test-set composed of patients’ of Mayo Clinic.

### Visualization

The designed graph based neural networks need to be explained in terms of weights assigned to edges to understand the learnt similarity between cases. Ying el al. proposed - ‘GNN explainer’ that learns concise subgraph structures consisting of nodes that play an important role in assigning output labels to a node^24^. We use the GNN explainer framework to plot sample nodes predicted as true and false positives by the models on both external and internal datasets using optimized graph models. Edge weights indicate the relative importance of each connection in predicting the label for the center node. We designed a dashboard with Panel 1 showing the GNN explainer similarity analysis. Panel 2 shows chest X-ray forming node features. Panel 3 shows an ordered list of common CPT subgroups in the neighborhood. Panel 4 shows demographic features of the patient corresponding to the center node.

## Results

### Quantitative performance

We report the performance of the two binary predictors (Model-1: Discharge from hospital, Model-2: Mortality) that are part of our framework, and comparative models in terms of class-wise and aggregated (weighted average) precision, recall, and F-score as well as confidence interval (95% confidence) on a randomly held-out set of test samples, in Table 2. Statistical significance of the difference in performance has been measured through p-values computed by statistical t-test with the best performing model as the reference. In Figure 3, we also represent the receiver operating characteristics (ROC) curves for these evaluations. Reported area under the ROC curve clearly indicates the superiority of the proposed graph-based models with edge structure based on ICD and CPT codes.

**Table 2.**
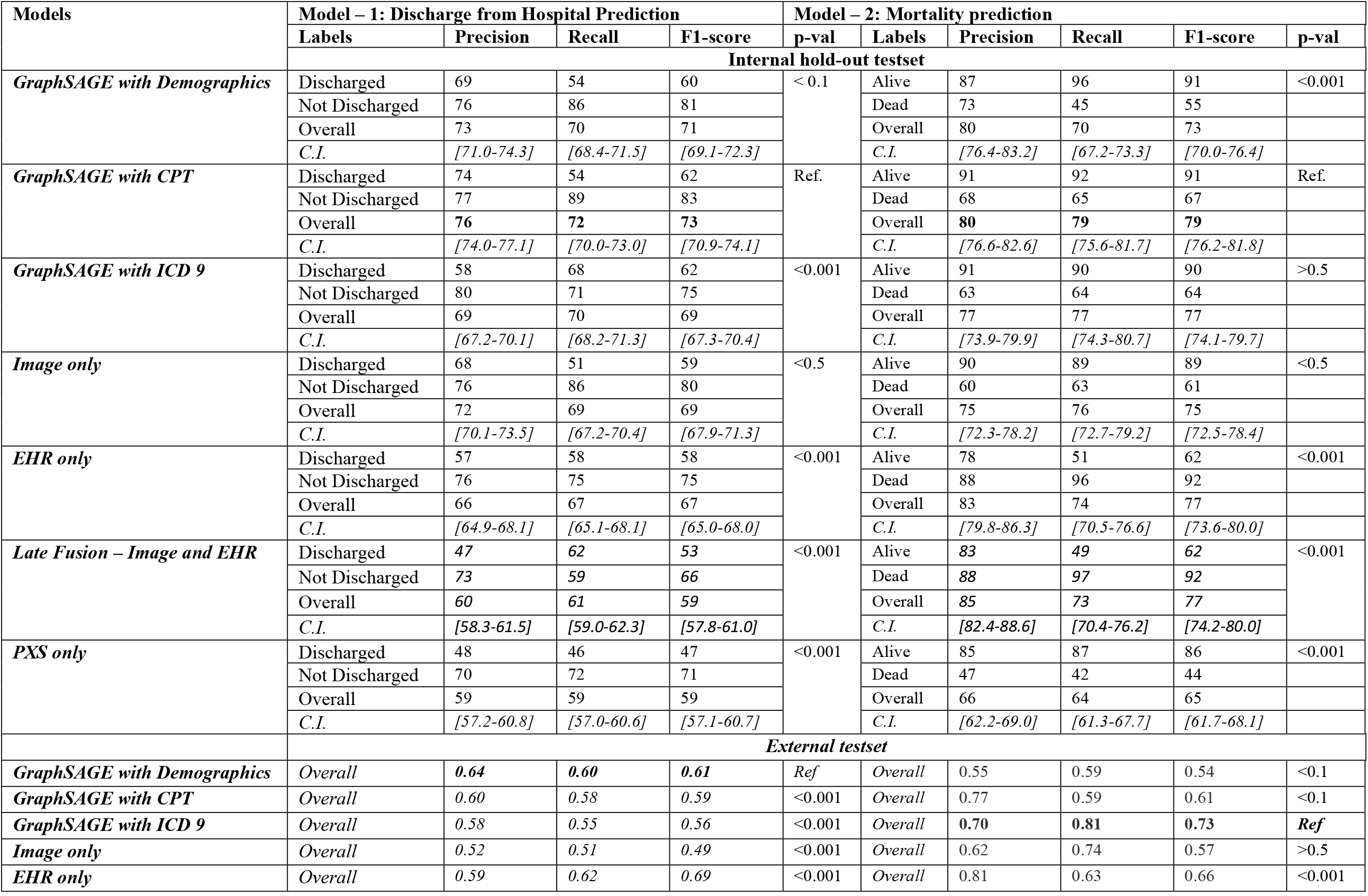
Performance of Model 1 – Prediction of discharge from hospital within 3 days and Model 2 – Mortality prediction on internal and external sets

**Figure 3.**
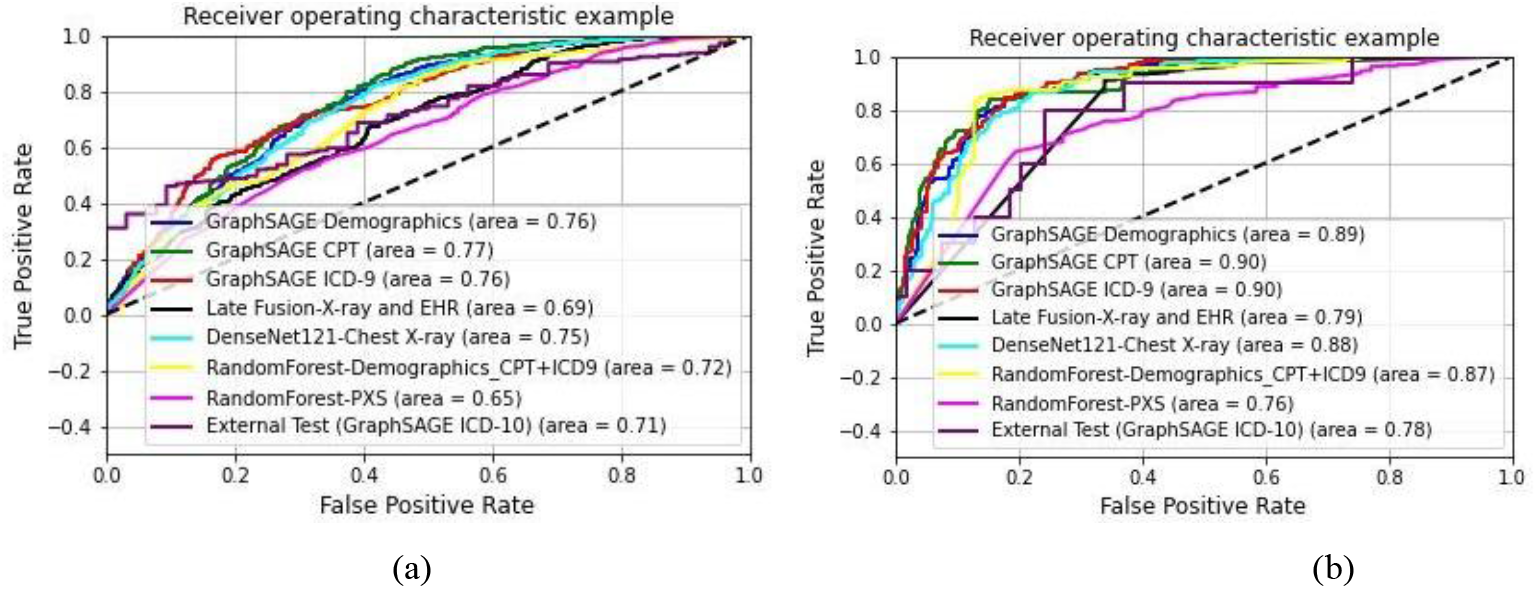
ROC curves; (a) Model-1: discharge prediction, (b) Model-2: Mortality Prediction. Each curve represents an individual model. Model names in the legends are formulated as - ‘Model Architecture-Input modality’. External performance only evaluated on the best model was included.

Table 2 show the performance of comparative predictive models - (1) *EHR only:* Random forest model using all EHR data sources as input (demographics, CPT groups, ICD-9 groups), (2) *PXS only:* Random forest model using only PXS score^23^ derived from the images as input, using a CNN model which was pretrained on ~160,000 images from CheXpert and fine tuned by transfer learning on 314 CXRs from patients with COVID-19, (3) *Image only*: softmax classification of the pretrained DensenET-121 using chest X-ray as input, (4) *Late Fusion*: fusion of label probability estimates from *EHR only* and *Image only* models, and (5) *GraphSAGE with XX:* GraphSAGE network with graph structure based on different EHR sources (XX) like demographics, CPT groups, and ICD-9 groups.

For both prediction of discharge from hospital and mortality, GraphSAGE with CPT and ICD achieved the optimal performance (no statistical difference), surpassing baseline single modality models - Image-only and EHR only, while PXS-only model also achieved suboptimal performance. Hence, clinical history seems more important for prediction of inpatient outcomes than demographic features. We experimented with combining ICD and CPT codes for graph formation. Such graph formation did not improve performance any further than what is achieved by these graphs based on ICD and CPT codes separately.

External validation was performed on data here the common comorbidities (diabetics, renal disease) are rare (<20%) compared to our internal training data (>55%) and as a consequence the respective procedures are less common. In such a distribution shift, often the machine learning model failed to generalize. This scenario provides an excellent opportunity to test the generalizability of graph based models. GCN training on internal data requires node features of the external data to be in the same space as node features of the internal data. There is no such strict requirement for edge features. They can be defined by any suitable similarity assessment. We employed common characteristics of external data to build edges for the external set and applied our GCN model trained on the internal data. The AUROC plot is shown in Fig 3.. It is clear that graph-based models have higher and more consistent generalization capabilities than baseline models (image-only and EHR-only) achieving better performance for both tasks.

### Qualitative evaluation

Results of the Dashboard exploration are shown in Figures 4 (Model 1) and 5 (Model 2). In panel view, panel 1 shows the ‘neighborhood’ of the central node with node colors indicating labels of nodes and edge thickness indicating importance of the connection/edge optimized by the predictive model for down-stream prediction tasks. Figures 4a-b show that the model is able to focus on a neighborhood with a large number of positive label nodes and assign appropriately high weights to their connecting edges while making true positive predictions. Figure 5a-b shows similar true positive predictions for Model-2 where the positive label indicates mortality within 3 days. Distribution of this label is highly imbalanced with positive labels as minority. In such a case, the model learns to predict a positive label even when only a few positively labeled nodes are present in the neighborhood. Given the scarcity of these positive data points, presence of even a few positively labeled nodes is considered as important evidence for graph-based learning.

**Figure 4.**
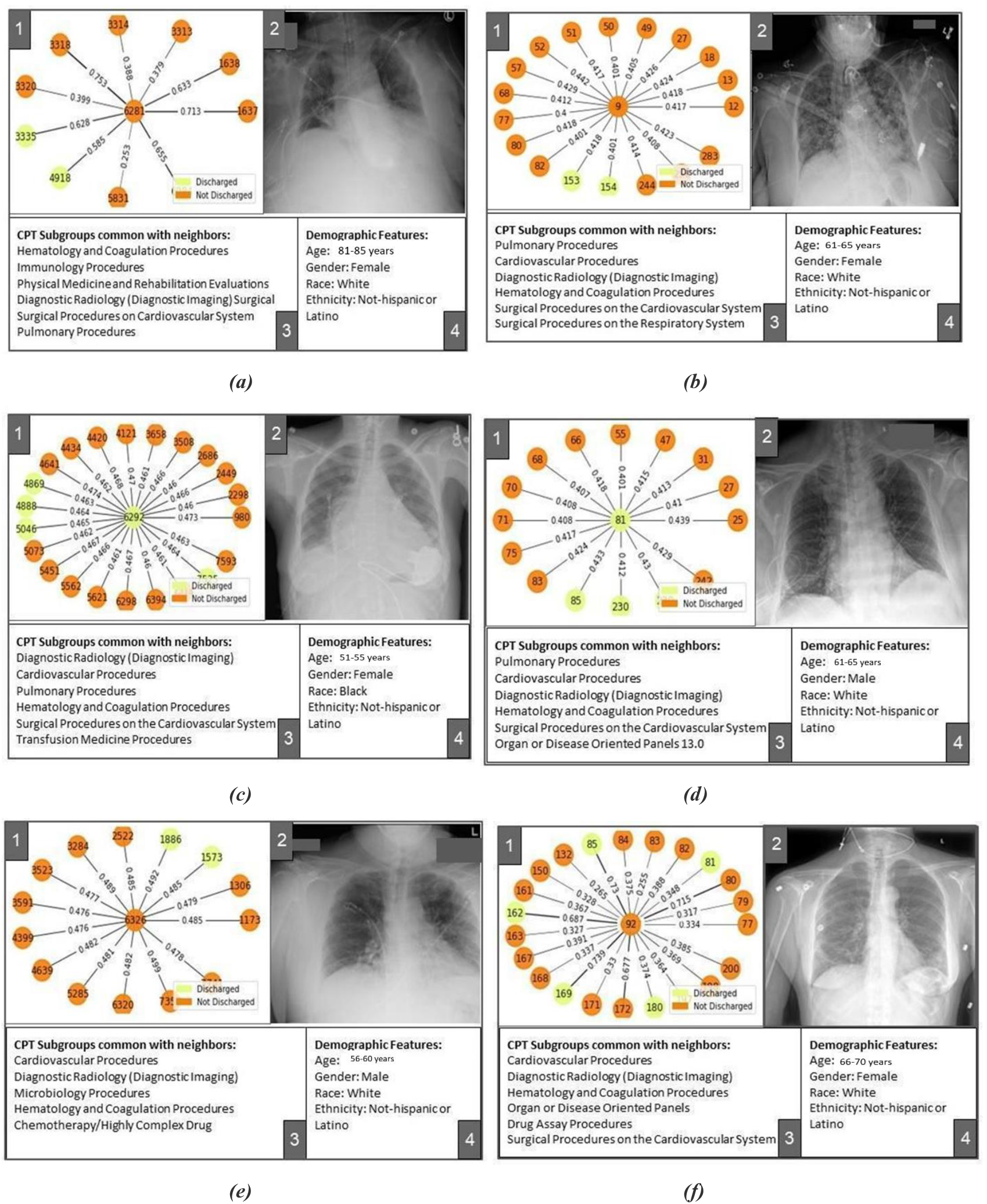
Pavel view of Model-1 (+ve label: Not Discharged in 3 days) results for internal and external sets results; (a): True positive prediction in internal set, (b): True positive prediction in external set, (c): False positive prediction in internal set, (d): False positive prediction in external set, (e): False negative prediction in internal set, (c): False negative prediction in external set

**Figure 5.**
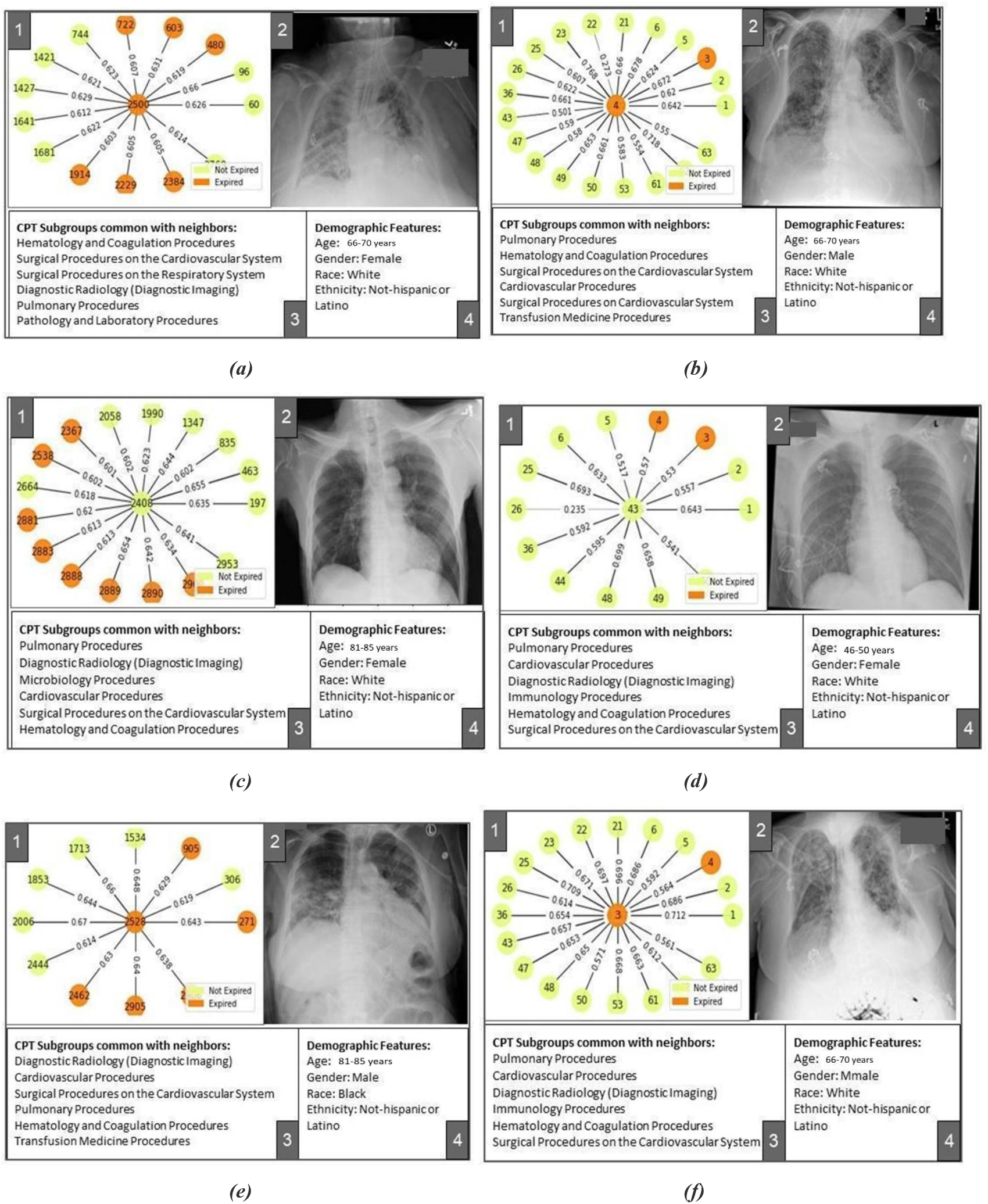
Pavel view of Model-2 (+ve label: Dead in 3 days) results for internal and external sets results; (a): True positive prediction in internal set, (b): True positive prediction in external set, (c): False positive prediction in internal set, (d): False positive prediction in external set, (e): False negative prediction in internal set, (c): False negative prediction in external set

While predicting adverse outcomes like mortality and stay in the hospital for more than 3 days, the model optimized a ‘neighborhood’ of patients formed due to all patients in the neighborhood undergoing cardiovascular and pulmonary procedures (surgical or non-surgical) as shown in panels 3 of Figures 4a-b and 5a-b. This is true for both the internal and external dataset while the standard of procedure coding is different in two institutions and thus the CPT subgroup distribution is significantly different for the two sets.

Patients corresponding to Figures 4a-b and 5a-b underwent mechanical ventilation categorized under the pulmonary procedures subgroup in the intensive care unit recorded in their history. Model optimized their representation in reference to a neighborhood of other patients undergoing pulmonary procedures (panels 3 of Figures 4a-b and 5a-b). This partially explains the model’s adverse predictions for all of these patients. Figures 4e-f show cases where the model falsely predicts patients to be discharged. Corresponding patients did not undergo ventilation assistive procedures under the pulmonary procedures subgroup. Model’s falsely favorable prediction can be explained by the neighborhoods optimized for these patients where the pulmonary procedure subgroup is not common (panels 3 of Figures 4e-f).

### Error Analysis

Dashboard based visualization helps us understand the limitations of graph-based modeling. Some of the common procedures (such as Diagnostic radiology procedures) form an easy way of connecting several patients together while it may lack predictive power. Patients corresponding to Figure 4c experienced a long stay in the hospital (16 days) and underwent ventilation assistance (Pulmonary Procedure subgroup) and several cardiovascular procedures. Model seemed to have focused on these factors while missing out on prediction of discharge from the hospital at the end of the long stay. Patient corresponding to Figure 5e was hospitalized for 4 weeks and went through several kidney disease related procedures. Due to the prevalence of cardiovascular and pulmonary procedures in the internal set, the model seems to have optimized a neighborhood where this patient shares cardiovascular and pulmonary procedures with other patients rather than kidney related procedures, thus mis-predicting the label for this patient. Such challenges can be addressed by including data temporality in the graph formation.

Figures 4c-d show cases where the model optimized a neighborhood full of positively labeled nodes while the central nodes were negatively labeled. Label distribution in the neighborhood may have affected the model’s judgment to falsely predict positive labels for central nodes. Figure 4e shows an interesting case where an optimized neighborhood has a large number of positively labeled nodes, and the model also learnt to put high emphasis on edges connecting such positively labeled nodes while the final prediction was still falsely negative. Corresponding normal chest X-ray in Panel 2 may explain the prediction of negative labels.

## Conclusion

We proposed a graph-based framework to preserve interdependencies in multi-modal data and similar cases to predict future clinical events (e.g. discharge and mortality) for the in-patients population tested positive for COVID-19 within 3 days of admission. During graph-based learning, in theory, two-fold information fusion of node features and graph structure ensure that relevant features (nodes and graph structure) for the targeted task are amplified while similar non-relevant attributes are suppressed. To our knowledge, this is the first attempt to encode and learn the patient-wise similarity within imaging data using a GCN model for predictive modeling for hospital resource optimization and interpretation using a dashboard. The proposed framework may face limitations in terms of application scope as it requires imaging data to be collected on a regular interval and was trained on data collected from highly integrated academic healthcare systems. Prediction interval is limited to 3 days which is still longer than most studies done in the past^25–26^.

## Data Availability

All data produced in the present study are available upon reasonable request to the authors

## Footnotes

1 adni.loni.usc.edu

2 https://fcon_1000.projects.nitrc.org/indi/abide/

3 https://www.aapc.com/codes/cpt-codes-range/

